# Plasma ACE2 activity is persistently elevated following SARS-CoV-2 infection: implications for COVID-19 pathogenesis and consequences

**DOI:** 10.1101/2020.10.06.20207514

**Authors:** Sheila K Patel, Jennifer A Juno, Wen Shi Lee, Kathleen M. Wragg, P. Mark Hogarth, Stephen J Kent, Louise M Burrell

## Abstract

COVID-19 causes persistent endothelial inflammation, lung and cardiovascular complications. SARS-CoV-2 utilises the catalytic site of full-length membrane-bound angiotensin converting enzyme 2 (ACE2) for cell entry causing downregulation of tissue ACE2. We reported downregulation of cardiac ACE2 is associated with increased plasma ACE2 activity. In this prospective observational study in recovered COVID-19 patients, we hypothesised that SARS-CoV-2 infection would be associated with shedding of ACE2 from cell membranes and increased plasma ACE2 activity.

**Methods:** We measured plasma ACE2 catalytic activity using a validated, sensitive quenched fluorescent substrate-based assay in a cohort of Australians aged ≥18 years (n=66) who had recovered from mild, moderate or severe SARS-CoV-2 infection (positive result by PCR testing) and age and gender matched uninfected controls (n=70). Serial samples were available in 23 recovered SARS-CoV-2 patients.

**Results:** Plasma ACE2 activity at a median of 35 days post-infection [interquartile range 30-38 days] was 97-fold higher in recovered SARS-CoV-2 patients compared to controls (5.8 [2-11.3] vs. 0.06 [0.02-2.2] pmol/min/ml, p<0.0001). There was a significant difference in plasma ACE2 activity according to disease severity (p=0.033), with severe COVID-19 associated with higher ACE2 activity compared to mild disease (p=0.027). Men (n=39) who were SARS-CoV-2 positive had higher median plasma ACE2 levels compared to women (n=27) (p<0.0001). We next analysed whether an elevated plasma ACE2 activity level persisted following SARS-CoV-2 infection in subjects with blood samples at 63 [56-65] and 114 [111-125] days post infection. Plasma ACE2 activity remained persistently elevated in almost all subjects, with no significant differences between timepoints in post-hoc comparisons (p>0.05).

**Discussion:** This is the first description that plasma ACE2 activity is elevated after COVID-19 infection, and the first with longitudinal data indicating plasma ACE2 activity remains elevated out to a median of 114 days post-infection. Larger studies are now needed to determine if persistent elevated plasma ACE2 activity identifies people at risk of prolonged illness following COVID-19.

## Background

COVID-19 causes persistent endothelial inflammation, lung, cardiovascular, kidney and neurological complications as well as thromboembolic phenomena of unclear pathogenesis [1]. SARS-CoV-2 utilises the catalytic site of full-length membrane-bound angiotensin converting enzyme 2 (ACE2) for cell entry [2] causing downregulation of tissue ACE2. We have previously reported that downregulation of cardiac ACE2 is associated with increased plasma levels of the catalytically active site of ACE2 [3]. In this prospective observational study in recovered COVID-19 patients, we hypothesised that SARS-CoV-2 infection would be associated with shedding of ACE2 from cell membranes leading to increased plasma ACE2 activity levels.

## Methods

We measured plasma ACE2 catalytic activity using a validated, sensitive quenched fluorescent substrate-based assay as previously described [3-5] in a cohort of Australians aged ≥18 years (n=66) who had recovered from mild, moderate or severe SARS-CoV-2 infection (positive result by PCR testing) and age and gender matched uninfected controls (n=70). Serial samples were available in 23 recovered SARS-CoV-2 patients. The immune responses in a subset of this cohort have been reported. [6] The study was approved by the University of Melbourne Human Research Ethics Committee (#2056689); all participants provided written informed consent in accordance with the Declaration of Helsinki.

Data were analysed with SPSS version 26.0. Plasma ACE2 activity levels were compared using the Mann-Whitney test or Kruskal–Wallis test and significance adjusted by Bonferroni correction for multiple tests. Serial ACE2 activity levels were analysed using the Friedman test for repeated measures; after post hoc analysis with Wilcoxon signed-rank tests with a Bonferroni correction applied. Two-tailed P-values <0.05 were considered significant.

## Results

The controls and SARS-CoV-2 recovered subjects were matched for age (54 ± 11 vs. 53 ± 14 years, p=0.49) and sex (37 (53%) vs. 39 (59%) male, p=0.46) (mean ± SD).

Plasma ACE2 activity at a median of 35 days post-infection [interquartile range 30-38 days] was 97-fold higher in recovered SARS-CoV-2 patients compared to controls (5.8 [2-11.3] vs. 0.06 [0.02-2.2] pmol/min/ml, p<0.0001, Figure 1A). There was a significant difference in plasma ACE2 activity according to disease severity (p=0.033), with severe COVID-19 associated with higher ACE2 activity compared to mild disease (11.2 [8.3-23.2] vs. 5.4 [1.8-9.0] pmol/min/ml (Figure 1B, p=0.027). Men (n=39) who were SARS-CoV-2 positive had higher median plasma ACE2 levels compared to women (n=27) (9.2 [5.8-15.3] vs. 2.1 [0.2-5.1] pmol/min/ml, p<0.0001), consistent with our previous findings in non-COVID-19 patients that males have higher plasma ACE2 activity levels than females [5].

**Figure 1:**
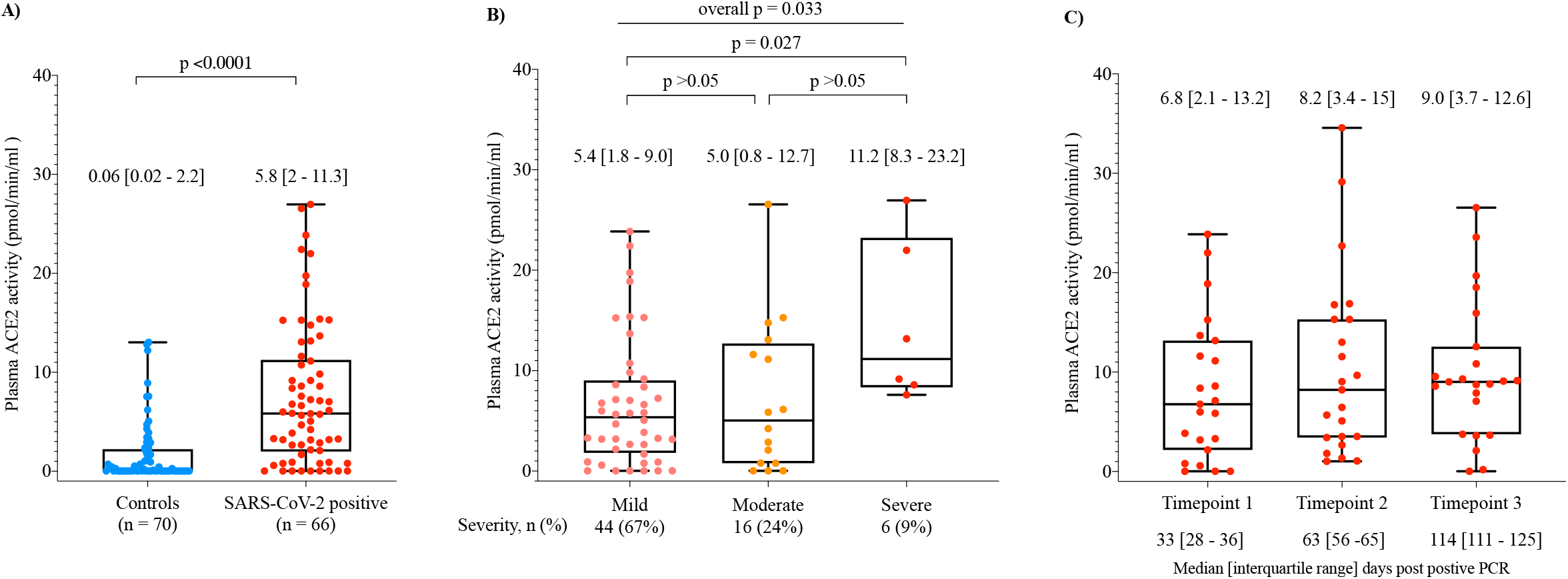
Plasma ACE2 activity is elevated in patients who recovered from SARS-CoV-2 infection (A), is increased with disease severity (B), and remains elevated 2-3 months after SARS-CoV-2 infection. Data shown as scatter points and box plot with whiskers representing min and max values of median plasma ACE2 activity between controls, and patients who recovered from SARS-CoV-2 infection (A), and median plasma ACE2 activity in patients recovered from SARS-CoV-2 infection according to disease severity (B). The median [interquartile range] plasma ACE2 activity levels are shown above each bar. The median plasma ACE2 activity in patients recovered from SARS-CoV-2 infection with serial samples (n=23) are shown in (C). The median [interquartile range] plasma ACE2 activity levels are shown above the bar for each timepoint. Two-tailed P-values <0.05 were considered significant.

We next analysed whether an elevated plasma ACE2 activity level persisted following SARS-CoV-2 infection in 23 subjects with blood samples at 63 [56-65] and 114 [111-125] days post infection. This subset was representative of the overall cohort for age (52 ± 15 years), gender (14 (61%) male) and disease severity (mild 15 (65%), moderate 5 (22%), severe 3 (13%). Plasma ACE2 activity remained persistently elevated in almost all subjects (Figure 1C), with no significant differences between timepoints in post-hoc comparisons (p>0.05). There was no change in plasma ACE2 activity in serial samples from a limited number of control subjects (n=8, timepoint 1: 0.8 [0.02-3.0] vs. timepoint 2: 1.4 [0.02-5.9] pmol/min/ml, p=0.31).

## Discussion

This is the first description that plasma ACE2 activity is elevated in patients after COVID-19 infection, and the first with longitudinal data to indicate that plasma ACE2 activity remains elevated out to a median of 114 days post-infection, the last time point sampled. It is already known that males have higher plasma ACE2 activity then females, but this is the first report that plasma ACE2 activity levels are increased in those with more severe infection. Although the pathophysiologic cause and consequences of elevated plasma ACE2 activity in the setting of COVID-19 are unknown, we reported that increased plasma ACE2 is an independent risk factor for major adverse cardiac events in patients with cardiac disease [3, 5]. Others have recently shown that increased concentration of ACE2 was associated with increased risk of major cardiovascular events in a large global population-based study [7].

In conclusion, we suggest that binding of SARS-CoV-2 to membrane ACE2 has at least 2 effects. First, it downregulates membrane ACE2 causing a dysregulated local renin angiotensin system (RAS) that favours inflammation and ongoing tissue damage secondary to excess angiotensin II. Second, the dysregulated local RAS is associated with prolonged shedding of the catalytically active site of ACE2 into the circulation.

In addition to the early multisystem effects of COVID-19, a proportion of patients experience prolonged ill-health [8]. There is growing concern over long COVID-19, how to define it and how to manage it. Larger studies are now needed to determine if persistent elevated plasma ACE2 activity identifies people at risk of prolonged illness following COVID-19 and to address whether therapeutic strategies directed at a dysregulated renin angiotensin system can reduce COVID-19 complications.

## Data Availability

Data will be available on reasonable request

## Acknowledgements

We thank the generous participation of the study subjects for providing samples. We thank Adam Wheatley, Jane Batten and Helen Kent for help with the COVID-19 cohort, and Ping Huang and Thomas McConville for assistance with the ACE2 activity assays.

This study was supported by the Victorian Government (SJK, JAJ), an Australian government Medical Research Future Fund award GNT2002073 (SJK, PMH), the ARC Centre of Excellence in Convergent Bio-Nano Science and Technology CE140100036 (SJK), NHMRC program grants APP1149990 (SJK) and APP 1055214 (LMB), project grant GNT1145303 (PMH), Medical Research Future Fund award GNT 1175865 (SKP, LMB). JAJ and SJK are supported by NHMRC fellowships.

